# The role of epidemic spreading in seizure dynamics and epilepsy surgery

**DOI:** 10.1101/2022.08.22.22279085

**Authors:** Ana. P. Millán, Elisabeth C.W. van Straaten, Cornelis J. Stam, Ida A. Nissen, Sander Idema, Johannes C. Baayen, Piet Van Mieghem, Arjan Hillebrand

## Abstract

Epilepsy surgery is the treatment of choice for drug-resistant epilepsy patients, but one in three patients continue to have seizures one year after surgery. In order to improve the chances of good outcomes, computational models of seizure dynamics are being integrated into surgical planning to simulate the effects of the planned surgeries. These modelling frameworks require several conceptual and methodological choices, as well as large amounts of patient-specific data, which hinders their clinical applicability. To address this problem, we considered the patient-specific brain network, derived from magnetoencephalography (MEG) recordings, and a simple epidemic spreading model as the dynamical basis for seizure propagation. This simple model was enough to reproduce the seizure propagation patterns derived from stereo-tactical electroencephalography recordings (SEEG) of all considered patients (*N* = 15), when the patients’ resected areas (RA) were used as the origin of epidemic spreading. The model yielded a more accurate fit for the seizure-free (SF, *N* = 11) than the non-SF (NSF) group and, even though the difference between the groups was not significant, the goodness-of-fit distinguished NSF from SF patients with an area under the curve AUC = 84.1%. We also explored the definition of a population model that combined data from different patients to fit the model parameters but was still individualized by considering the patient-specific MEG network. Even though the goodness-of-fit decreased compared to the individualized models, the difference between the SF and NSF groups held, and in fact became stronger and significant (*p* = 0.023), and the group classification also improved slightly (AUC= 88.6%). Therefore, combining data from different patients may pave the way not only to generalize this framework to patients without SEEG recordings, but also to reduce the risk of over-fitting and improve the stability of the models. Finally, we considered the individualized models to derive alternative hypothesis of the seizure onset zones and to test the surgical strategy *in silico* for each patient. We found that RA regions were on average more likely to originate the seizures, but that alternative explanations were possible. Virtual resections of the RA when considering these alternative seeds significantly reduced seizure propagation, and to a greater extend for SF than NSF patients (although the difference was not significant). Overall, our findings indicate that spreading models based on the patient-specific MEG network can be used to predict surgical outcomes, with better fit results and greater reduction on seizure spreading linked to higher likelihood of seizure freedom after surgery.

## 1 Introduction

Epilepsy is a highly prevalent neurological disorder, affecting between 4 and 10 per 1000 people worldwide [1]. About 1 out of 3 people who suffer from epilepsy do not respond to medication, i.e. they present drug-resistant or refractory epilepsy [2]. In these cases, epilepsy surgery (ES), consisting of the removal or disconnection of the necessary brain regions to stop seizure propagation -/namely the *epileptogenic zone* [3]– is the treatment of choice. Several conditions must be met for the surgery to proceed, including that a focal origin of the seizures can be found, and that the proposed surgery can be performed safely, i.e. without unwanted side-effects such as sensorimotor deficits, amnesia, or aphasia. Surgery outcomes vary greatly depending on epilepsy type, with seizure freedom attained for about 2*/*3s of the patients one year after surgery [4]. Although the majority of patients still experience a reduction in seizure frequency or intensity after surgery, even when surgery is not completely successful, side-effects and cognitive complaints are also common, and can be difficult to predict accurately on an individual basis [5].

In recent years, several efforts have been made to improve the outcome of epilepsy surgery. One important conceptual leap forward is the notion of *epileptogenic networks* [6], according to which even in case of focal epilepsy the *epileptogenic focus* should not be considered as solely responsible for seizure generation or propagation, but rather the existing brain network also plays a role in promoting (or inhibiting) the ictal state [7–9]. Within this perspective it has been found that several properties of the brain networks of epilepsy patients deviate from those of healthy controls [10–14]. In particular, abnormalities are often found relating to the *brain network hubs*, which often suffer from targeted damage in patients with neurological disorders (see Stam [8] for a review). In the case of epilepsy, hubs may facilitate the propagation of epileptiform activity throughout the brain [15, 16]. In fact, several studies have pointed out the existence of *pathological hubs*: abnormal, hyperconnected regions in the vicinity of the epileptic focus, which mediate seizure propagation [15, 17–19].

Within the network perspective, the effect of surgery is no longer straightforward to predict: local changes in a network may have widespread effects, or be compensated by the remaining network [20, 21]. Moreover, the specific effect of a surgery will depend on the individual network configuration [22], making it fundamental to consider patient-specific connectivity in order to tailor the surgery specifically to each patient. Network-based studies have found group-level differences between seizure-free and non-seizure free patients [19, 21, 23], for instance Nissen et al. [18] found that the removal or a pathological hub, or a region highly connected to it, was strongly associated with seizure freedom.

A data-driven manner to address this problem is via *computational models of epilepsy surgery*, which simulate *in silico* different resection strategies to help predict their impact before hand, with the goal of improving the planning of resective epilepsy surgery [24–33]. In order to tailor the resection strategy for each patient, and thus increase the chances of seizure freedom, the models are fitted to patient-specific data such as the underlying brain network connectivity (derived via different imaging techniques), stereo-typical patterns of seizure propagation and clinical biomarkers of the suspected location of the epileptogenic focus. Once the models have been defined, they can be used to predict the outcome of surgery [30, 31], or to propose alternative resection strategies, for instance in the case of a previous bad outcome or inoperable regions [31], or with a smaller impact than the actual surgery [25, 33–35].

The computational models of epilepsy surgery rely on the definition of a dynamical model of seizure generation and propagation. However, the specific mechanisms underlying seizure dynamics are not well known, and likely not unique: epilepsy is a heterogeneous disorder, and at least 6 different stereotypical patterns of seizure dynamics have been distinguished [24, 36, 37]. Thus, assumptions must be made in the modeling of seizure dynamics, and different levels of description, at different scales, are possible [38]. Realistic models make use of highly detailed non-linear dynamics [39], such as population-rate models [40] or neural-mass models, combined with one or several slow variables to account for the transition from normal to ictal activity [26, 30]. Within this perspective, several studies have tried to model seizure dynamics and predict the outcome of epilepsy surgery, with remarkable success at a group level: Sinha et al. [31], using a dynamical model based on EEG connectivity to identify epileptogenic regions, found that the overlap between these regions and the RA predicted surgery outcome with 81.3% accuracy. Proix et al. [37] found that their seizure model, the *epileptor model* [41], defined over MRI networks, could distinguish between good (Engel class I) and bad (Engel class III) outcomes. Further studies within this modelling framework also found a better match between the hypothesized EZ and propagation zone (i.e. the first regions to which ictal activity propagates to) for SF than NSF patients [42, 43]. On a virtual resection study, Sip et al. [44] found that the effect of the resection in the model correlated with surgical outcome, so that patients with Engel score I and II presented a significantly larger effect of virtual resections in the model. Finally, Goodfellow et al. [28] also found significant differences in the model prediction for Engel Class I and class IV patients, using an electrocorticogram (ECoG) modelling framework.

Detailed models of ictal activity, however, come at a high cost: several parameters need to be adjusted beforehand, with unavoidable arbitrary choices. This complicates the setting of the model parameters and either large quantities of data are needed, or several assumptions must be made. As a consequence there is a high risk of over-fitting, and generalizing the results to new data-sets becomes troublesome. In order to solve this problem, in-depth studies to characterize the dynamical properties of the models, and the interplay between network structure and emergent dynamics, are needed [45–47], often in combination with elaborate modelling optimization frameworks, such as Bayesian inference [43, 44, 48, 49] or deep learning [50]. Another possibility to circumvent these issues is by considering simpler, abstract models that focus only on the behavior of interest: the propagation of ictal activity throughout the brain [33, 34], accounted for by the slow permittivity variable of the epileptor and similar highly dimensional models Sip et al. [44]. Conceptually, this process is equivalent to other spreading processes on networks, a problem that has been well-characterized by means of *epidemic spreading models* [51]. Epidemic spreading models simulate the propagation of an agent from some given location on a network to other connected areas, a basic phenomenon appearing in a multitude of systems. In the case of brain dynamics, such models have been used to study the spreading of pathological proteins on brain networks [52, 53], or the relation between brain structure and function [54]. Due to their ubiquity and relative mathematical simplicity, epidemic spreading models are supported by a wealth of mathematical background characterizing the emergent dynamics in relation to different properties of the underlying network. This information can later be useful for clinical applications, e.g. general rules for spreading phenomena on complex networks that can be applied to understand seizure propagation.

In previous studies we considered epidemic spreading models as the basis for seizure propagation over the brain, without trying to mimic the complicated biophysical mechanisms involved in the process [33, 34]. Within this framework, we found that epidemic spreading models fitted with patient-specific data could reproduce the stereotypical patterns of seizure propagation on patient-specific brain networks, individually for each patient. Moreover, by taking into account this patient-specific connectivity, alternative or smaller resections could be found with the model, which we hypothesized could lead to fewer side effects with the same outcome, in terms of seizure reduction [33, 34].

Here we consider an epidemic spreading model to generate individualized seizure propagation models that are based on the patient-specific MEG connectivity and seizure propagation pathways as derived from invasive EEG recordings. This framework generalizes on previous studies by our group [33, 34] by including a recovery mechanism in the spreading model, allowing the return to the healthy (post-ictal) state, so that seizures may remain local (i.e. if the affected regions recover before spreading the ictal state to distant regions) or generalize. We considered a group of 15 epilepsy patients who underwent epilepsy surgery, and for whom the surgical outcome at least one year after surgery was known. We illustrate how the present model can be used to generate alternative hypotheses on the seizure onset zone and test different resection strategies, and we discuss the challenges associated to the model fitting, even in this simple scenario, and associated risks. We also present a population model that integrates spreading data from all patients but can still be individualized and applied to patients without SEEG recordings. We discuss how this approach can help reducing the over-fitting risk and noise effects, and how it may increase the generizability and clinical application of epilepsy surgery models.

## 2 Results

The individualized seizure propagation models were based on an epidemic spreading model –the Susceptible – Infected - Recovered or SIR model– equipped with patient-specific data, as depicted in figure 1. A total of 15 patients (9 females) were included in the study, 11 of whom were seizure free (SF) one year after surgery (Engel Class 1A, see table 2 for the patient details).

**Figure 1:**
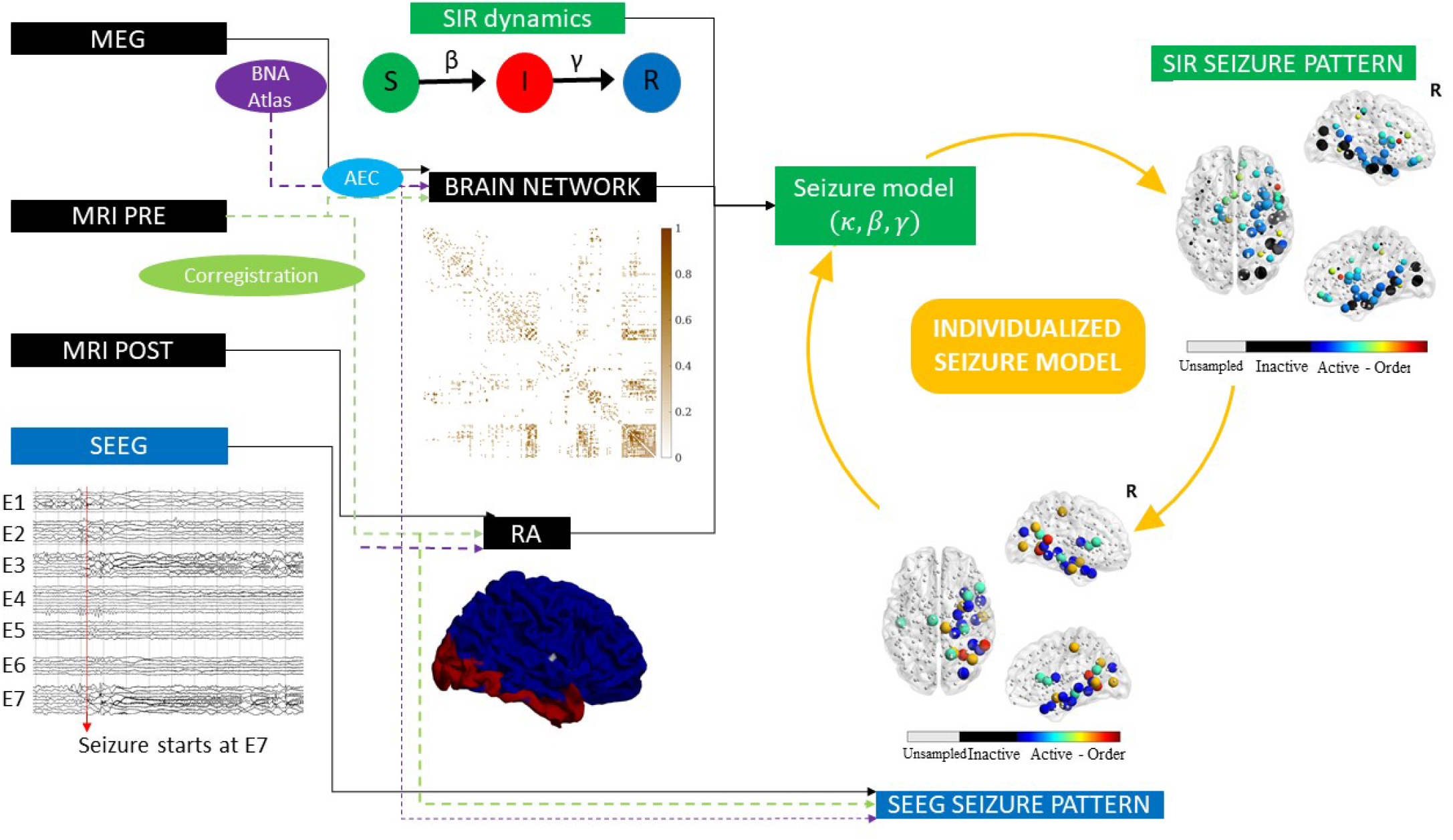
Sketch of the methodology followed in this study. The SIR model was used to simulate seizure propagation. As the backbone for the model dynamics, we used the patient-specific AEC-MEG network, and the seed regions were initially defined as the resection area (RA), which was reconstructed from the pre- and post-surgery MRIs. By analyzing the seizures generated by the model, we derived the *SIR seizure pattern*, describing seizure propagation in the model. This was compared to the *SEEG seizure pattern* as derived from SEEG recordings of ictal activity. The spreading patterns describe the activation order the active (i.e. infected, in the ictal state) and sampled (by the SEEG electrodes) ROIs. Comparison between the model and the data (see figure 2) allowed us to fit the model parameters to the SEEG pattern and create an *individualized seizure propagation model* for each patient.

### 2.1 Seizure propagation as an epidemic spreading process

Seizure propagation was modelled using the SIR model such that the susceptible (S), infected (I) and recovered (R) states accounted respectively for the healthy (pre-ictal), ictal and healthy (post-ictal) states. The SIR model describes the spreading of an epidemic process on a network from a set of seed regions to the other nodes, and it has been applied in a multitude of scenarios involving spreading phenomena. The emerging behavior of the system under this dynamics is well-characterized in relation to the underlying network structure [51, 55]. In this scenario, the model does not try to mimic the detailed biophysical processes involved in seizure generation and propagation, instead it is used here as an abstraction that includes only the most relevant features of seizure propagation [33, 34, 44, 51]. The model is characterized by two control parameters, the global spreading rate *β* characterizing the probability of spreading of the infected state, and the recovery rate *γ* characterizing the recovery probability of each infected node. The model was simulated on top of the patient’s brain network reconstructed from resting-state MEG recordings using the Brainnetome Atlas (246 nodes). Each region of interest (ROI) was represented via a *node i* in the network, and each connection via a *link* (*i, j*), with the weight *w*_*ij*_ of link (*i, j*) indicating the strength of the coupling between ROIs *i* and *j*. The weight distribution affected the spreading pattern as *w*_*ij*_ modulated locally the spreading rate: the probability that an infected node *i* infected a neighbour *j* was given by *βw*_*ij*_. Thus, strongly connected neighbours were more likely to propagate the infected state. As coupling metric we considered the uncorrected AEC (Amplitude Envelope Correlation). AEC-MEG networks include both short- and long-range functional connections, combining in one network aspects of structural and functional connectivity [34]. The networks were thresholded (but not binearized), with the link density *κ* acting as the third control parameter of the model. An exemplary case of the final weight matrix is shown in figure 1.

### 2.2 Individualized seizure propagation models

The seizure propagation model was adapted individually for each patient by fitting the simulated propagation patterns to patient-specific seizure propagation data derived from SEEG recordings and by setting the seed of epidemic spreading as the resected area (RA) (see 5.5.2 for more details). Two seizure propagation patterns were constructed, the *SIR* and the *SEEG seizure patterns*, depicting respectively the activation order of the sampled ROIs in the SIR- and SEEG-derived seizures. An exemplary case is shown in figure 1). The total correlation between the two patterns, *C*, as defined by Eq. 2 in the Methods, and illustrated in figure 2 (for the same exemplary case as in figure 1), was used as the *goodness of fit* of the model (see sec. 5.5.2).

**Figure 2:**
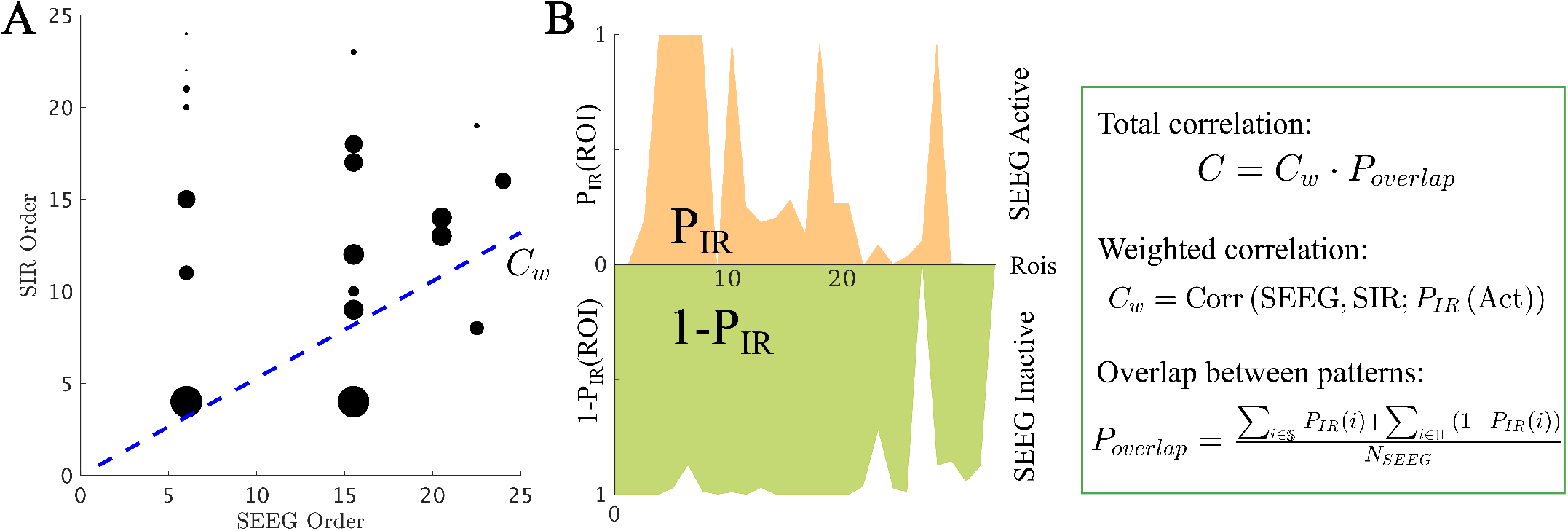
Total correlation *C* between the SIR and SEEG seizure patterns. First, the set of active (i.e. infected) ROIs in both patterns was identified, and the weighted correlation *C*_*w*_ between the activation orders was calculated (panel A, blue line). As correlation weights we used the probability that the ROI *i* was infected in the SIR pattern, *P*_*IR*_(*i*), depicted in the figure by the size of the black circles. Then, to control for the extension of the seizure in both patterns, we computed the weighted overlap between the active and inactive regions (panel B), *P*_*overlap*_. *P*_*IR*_(*i*) is the probability that the ROI *i* becomes infected during the spreading process. Conversely, 1 −*P*_*IR*_(*i*) is the probability that it does not become infected. S and H stand respectively for the sets of ROIs that are infected (i.e. in the seizure state) and not infected (i.e. in the healthy state) in the SEEG pattern. The total correlation was then defined as *C* = *C*_*w*_*P*_*overlap*_. For more details see section 5.5.2. The spreading patterns corresponding to this data are shown in figure 1 under “SIR Seizure Pattern” and “SEEG Seizure Pattern”, respectively.

Within this framework, we found the set of parameters (*κ, β, γ*) yielding the best model fit *C* for each patient (see Methods for details and figure 3A for the fit results). On average, we obtained a model fit of *C* = 0.30 (with standard deviation std_*C*_ = 0.20). The model provided a (not significantly) better fit for the SF (*C*_*SF*_ = 0.35) than NSF (*C*_NSF_ = 0.15) patient groups (*C*_*SF*_ − *C*_NSF_ = 0.20, *t*(13) = 1.79, *p* = 0.097, unpaired t-test), as shown in figure 3B. A ROC classification analysis based on the goodness of fit returned a good classification result with an area under the curve (*AUC*) of *AUC* = 0.841. There were no significant differences in the fit parameters between the groups (average ± standard deviation: *κ* = 25 ± 22, *β* = 0.02 ± 0.03, *γ* = 0.03 ± 0.04).

**Figure 3:**
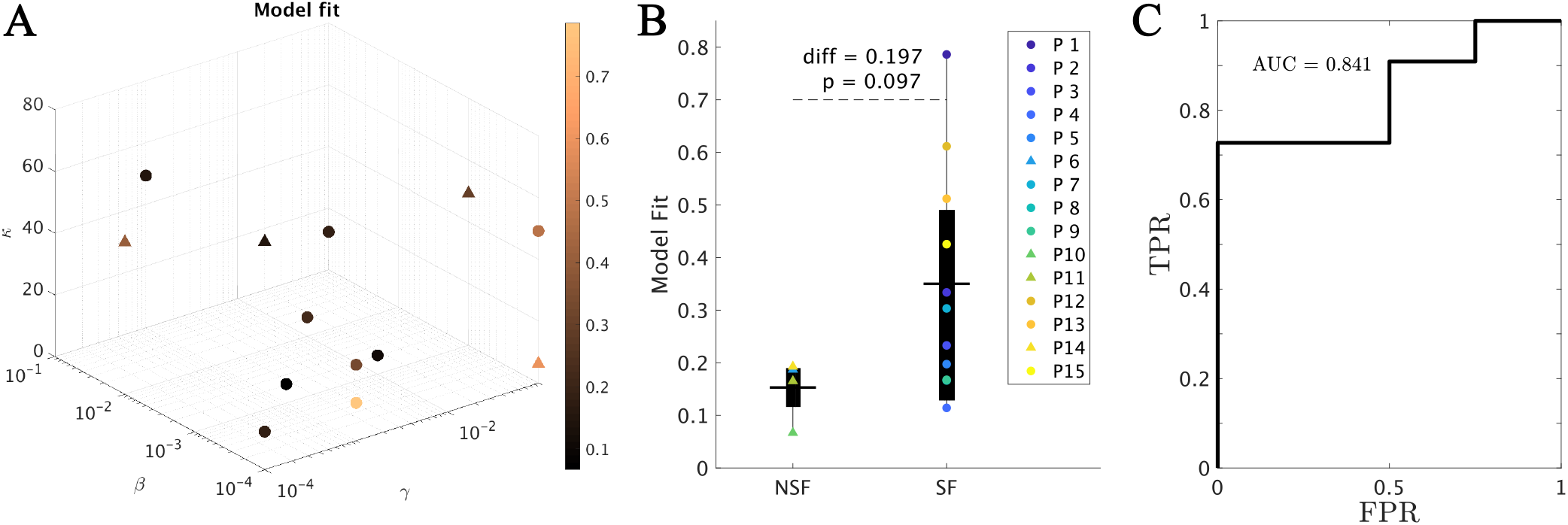
Model fitting results. **A** Best fit found for each patient. The model parameters (*κ, β, γ*) form the three axes and the color-code indicates the goodness of fit *C*. Note that some points overlap. **B** Group comparison of the goodness of fit, for the NSF and SF groups (unpaired t-test). Each color represents a different patient, as indicated by the legend. The solid lines on each box indicate the mean values. In panels A and B, SF (NSF) patients are indicated by circles (triangles). **C** ROC curve corresponding to the group classification according to the goodness of fit. A positive result was defined as a good outcome (SF). FPR indicates the false positive rate (NSF patients classified as SF), and TPR the true positive rate (SF patients classified as SF).

### 2.3 Population model

The population model 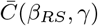 was defined by measuring the average fit across patients, and by re-scaling the spreading rate *β*_*RS*_ to combine in a single quantity the main parameters controlling the expected number of infected nodes, i.e. the original spreading rate, the density of connections (as given by *κ*) and the size of the seed. Details of this procedure are given in the methods (section 5.5.3). The resulting fit diagram (figure 4A) resembles a familiar phase transition diagram, with an interface of high goodness of fit (yellow regions) corresponding to a roughly constant spreading-to-recovery ratio *β*_*RS*_*/γ* = const. Most individual best fits (black markers) fell within this region, although there was large variability among the individual results (in fact, we found low signal to noise ratios of aprox. 1*/*5 as shown in the Supp. Information, Supp. Fig. S2).

**Figure 4:**
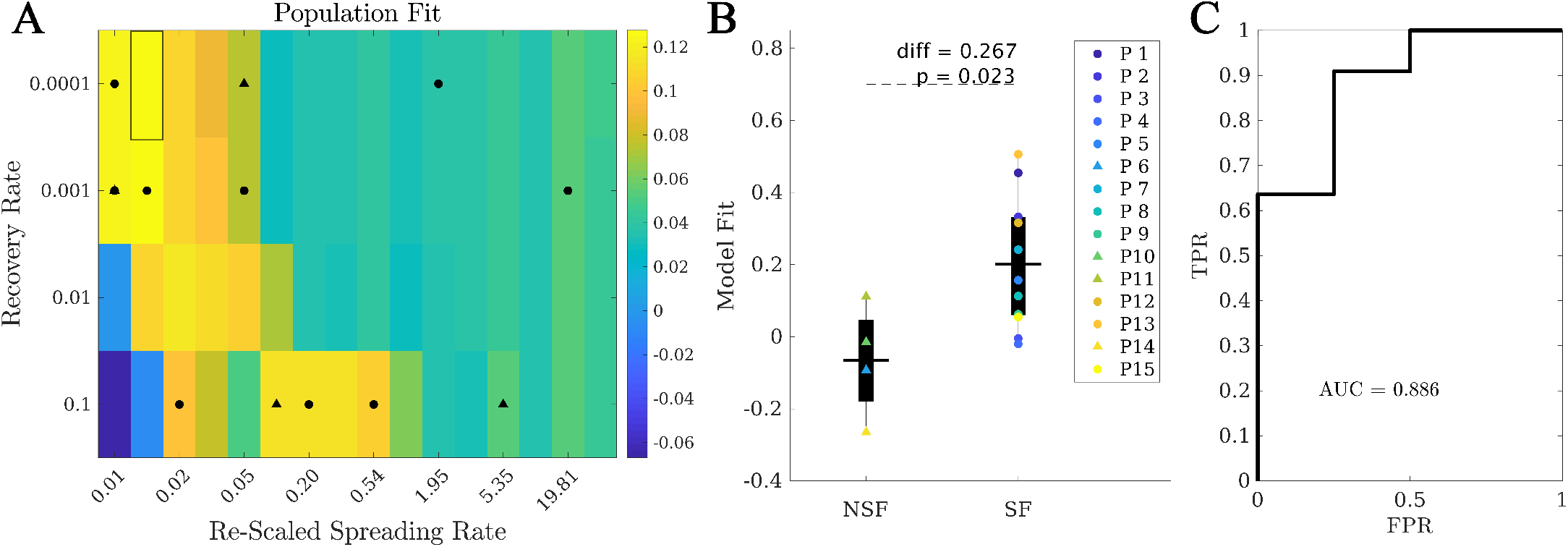
Population model. **A** Phase diagram showing the population fit 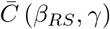. Black markers indicate the location of the best individual fits in this diagram, with circles (triangles) corresponding to SF (NSF) patients. The highlighted rectangle indicates the best fit. **B** Comparison between the population fit for the SF and NSF groups. Solid lines indicate the mean values for each group. **C** ROC classification analysis between the SF and NSG groups, with an AUC = 0.886.

Within this population model, the best fit was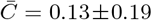, corresponding to *β*_*RS*_ = 0.01, *γ* = 10^−4^ (highlighted rectangle in figure 4). Remarkably, when considering the fit results for each patient at the optimal population point, we found that the SF group 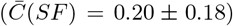 presented a significantly better fit than the NSF group 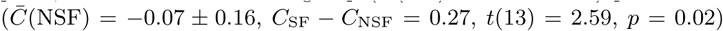, as shown in figure 4B. Moreover, the ROC classification analysis in this case also provided a good classification (AUC = 0.886) between the SF and NSF groups (see figure 4C).

### 2.4 Alternative Seizure Onset Zones

Once the model was fitted to the patient-specific seizure propagation patterns, we estimated the likelihood of each individual ROI acting as the SOZ, as given by the total correlation metric, *C*_*R*_, when ROI *R* was used as the single seed for the SIR dynamics. For this we considered the individualized spreading models (i.e. fitted individually for each patient) as they provided a better characterization of the individual SEEG spreading patterns. For all patients, single seeds could be found that provided a good approximation to the seizure propagation pathways, i.e. with high *C*_*R*_ values, as shown in figure 5 (left panels) for two exemplary cases. The *seed likelihood maps* depicted different degrees of localization and sparseness for different patients, as well as different degrees of overlap with the RA (also shown in figure 5 for comparison purposes, middle panels). From visual inspection, the RA tended to appear in regions with relatively high *C*_*R*_, but did not include the maximum. In order to test whether RA ROIs tended to have higher *C*_*R*_ than non-RA ROIs, we compared the seed likelihood for the two ROI sets, for each patient, as shown in the right-side panels in figure 5. In these two exemplary cases, RA ROIs were significantly more likely to be the seed than non-RA ROIs. However, this was the case only for 7 out of 15 cases, of which 1 was NSF. For the remaining 8 cases, no significant difference between the groups was found (see Supp. Table S1).

**Figure 5:**
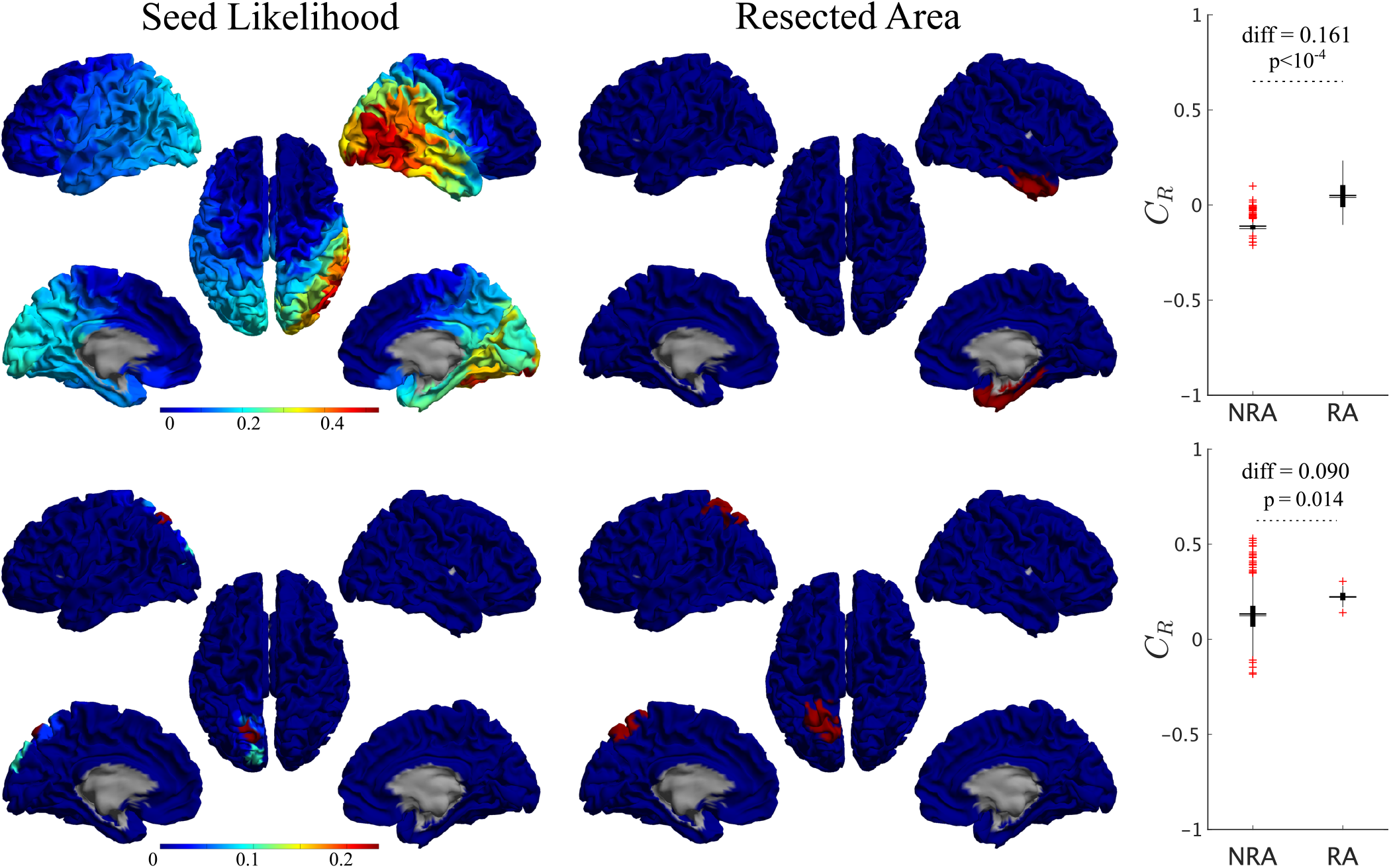
Seed-likelihood maps for two representative cases. Left panels show the seed-likelihood of each ROI, *C*_*R*_, for two exemplary cases (cases 13 and 15, respectively from top to bottom), whereas the middle panels show the corresponding resected areas in red. The right panels indicate the comparison between RA and non-RA (NRA) ROIs, for these two cases (unpaired t-test). The solid lines stand for the mean values. Red pluses mark outliers (1.5 times over the interquartile range).

At a group level, we found that RA ROIs were on average more likely to be the seed than non-RA ROIs, as shown in figure 6A (*C*_RA_ − *C*_NRA_ = 0.077, *p* = 0.016, *t*(14) = 2.74, paired t-test). However, the ROI with the maximum likelihood, *C*_best_, did not belong to the RA for any case (see for instance the two exemplary cases shown in figure 5). Thus, the most likely single seeds were close to the RA, but did not belong to it. Despite the individual best seeds (*Best*) performing better than the RA, the difference was not significant. Moreover, both the *Best* and *RA* seeds performed better than the averaged individual RA ROIs, ⟨*RA*⟩, and than random seeds of the same size as the RA, *RND* (see Supp. Information for details of the comparisons).

**Figure 6:**
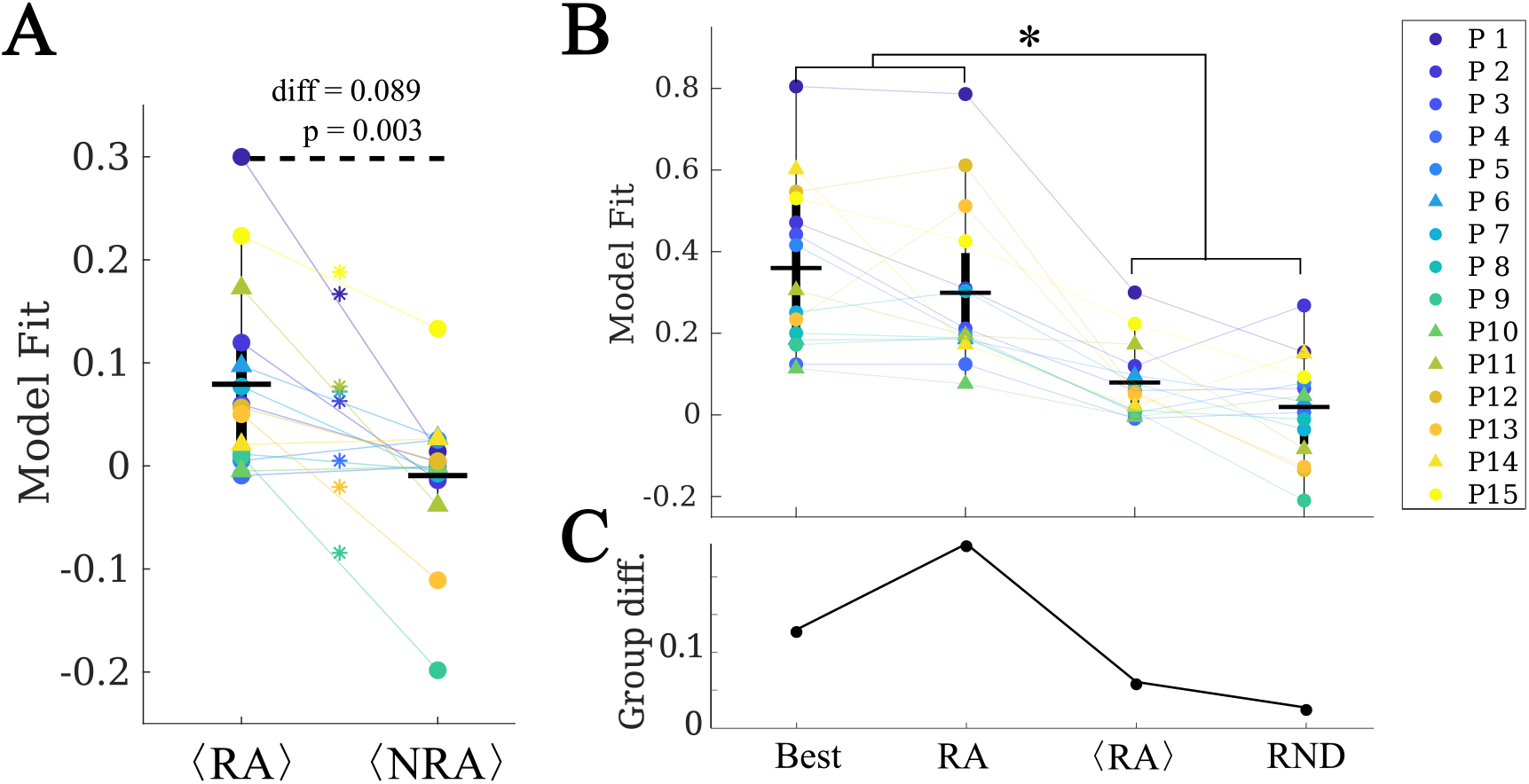
Analysis of alternative seeds. **A** RA regions had a significantly higher model fit (given by the total correlation when considering each ROI as the single epidemic seed, *C*_*R*_) than non-RA regions (NRA, paired t-test) on average. Stars indicate a significance difference at the patient-level. **B** Effect of the seed choice on the model fit. *Best* stands for the best single seed, *RA* for using the whole RA, ⟨*RA*⟩ for the average of the RA ROIs single seed fits, and *RND* for random seeds of the same size as the RA. **C** Group difference (SF vs NSF) found for each of the seed fits. The two groups only differed significantly when using the RA as seed.

No difference in the average seed-likelihood of the RA was found between the SF and NSF patients (*C*_RA,SF_ − *C*_RA,NSF_ = −0.006, *t*(13) = −0.09, *p* = 0.93, unpaired t-test), or in the maximum single seed-likelihood, *C*_max_ (*C*_best,SF_ − *C*_best,NSF_ = 0.08, *t*(14) = 0.6, *p* = 0.5), as shown in figure 6C. Moreover, the difference between the SF and NSF groups also vanished when considering random seeds (*C*_RND,SF_ − *C*_RND,NSF_ = −0.02, *t*(14) = −0.3, *p* = 0.8, figure 6).

### 2.5 Virtual resection analysis

We performed a virtual resection analysis to simulate the effect of the surgery for each patient, considering optimal seeds of increasing sizes (derived with a recursive procedure) and virtual resections of the RA (see the Methods section for details). For each patient, there was a significant decrease in seizure propagation with the surgery for all considered seed sizes, as given by the normalized decrease in spreading *δ*_VR_. We found that, on average, the SF group presented larger *δ*_VR_ (see figure 7A) for all considered sizes, but the difference was not significant in any case (see Supp. Fig. S2 for details of the comparisons). Finally, when considering the normalized decrease as a classification metric for SF versus NSF patients we found AUC values between 0.636 and 0.750 (average = 0.691), as shown in figure 7B.

**Figure 7:**
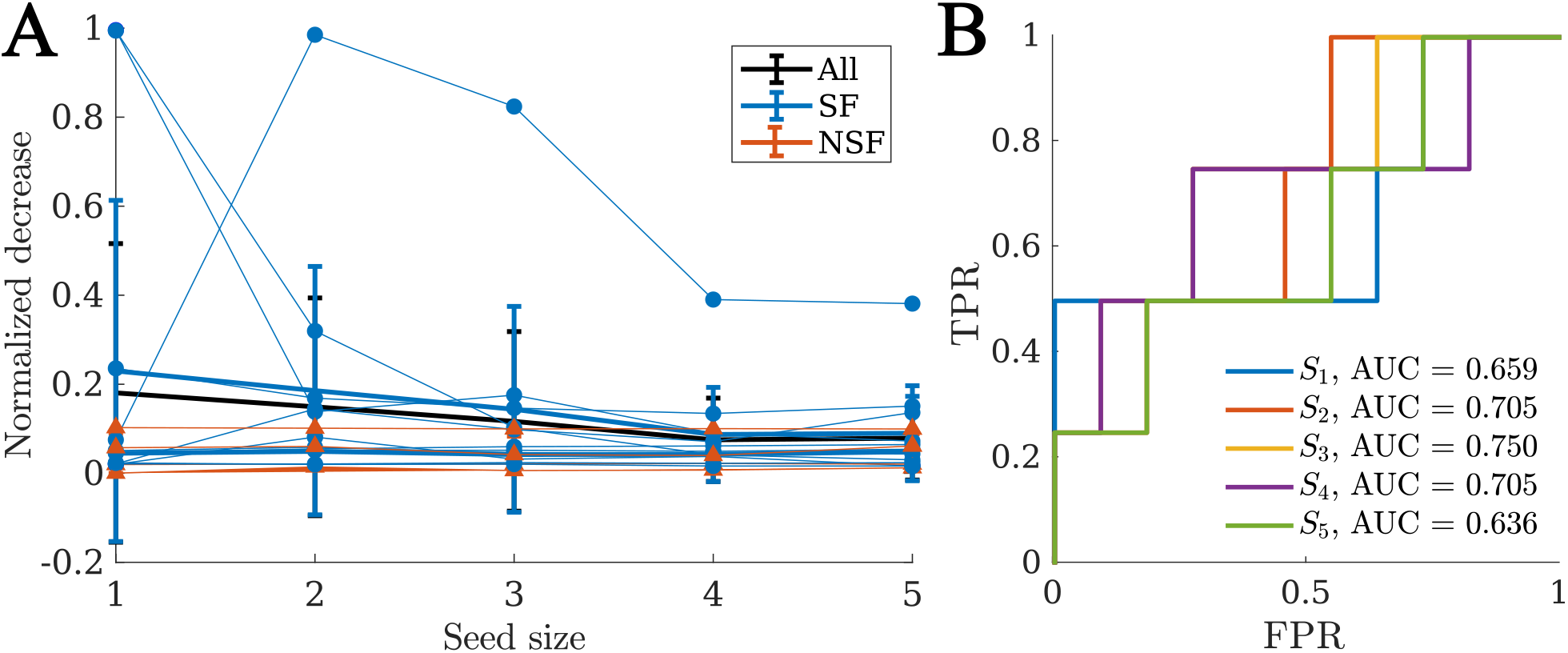
Virtual resection analysis. **A** Normalized decrease in spreading *δ*_VR_ for seeds of increasing sizes. Each data point indicates an individual patient, blue circles stand for SF patients and red triangles for NSF patients. The thick lines indicate the average values for the whole group (black, “All”), the SF (blue) and NSF (red) groups. **B** ROC classification analysis for SF versus NSF outcome, for each considered seed size as indicated in the legend.

In order to understand what defines the effect of the resection, we computed the correlation between the normalized decrease in logarithmic scale log (*δ*_VR_) and different dynamical and network properties (see table 1) that characterize the network structure before and after the resection, as well as the effect of the resection. We found that the largest amount of variance was explained by the size of the resection *S*_RA_, with larger resections leading to a larger effect of the VR, as one might expect. The centrality of the RA (given by the out-connectivity *E*_RA_ and betweenness centrality BC_RA_) also correlated significantly with the effect of the resection, although the effect was weaker. Remarkably, the baseline centrality properties of the seed regions (i.e. size *S*_seed_, out-connectivity *E*_seed_ and BC, BC_seed_) did not show significant effects, but the properties of the seed *after* the resection did show a significant negative correlation with the effect of the resection on spreading. The structural effect of the resection, given by the decrease in centrality of the seed (both out-connectivity Δ*E*_seed_ and BC ΔBC_seed_) was significantly and positively correlated with the dynamical effect. The model parameters also played a role in the effect of the virtual resections, with larger spreading-to-recovery ratios associated with larger effects of the surgery.

**Table 1:** Relation between the effect of VRs of the RA (as given by the normalized decrease in spreading after the resection) and the properties of the model, baseline and post-resection networks. As model parameter we consider the *spreading-to-recovery ratio βκ/γ*, which combines the three model parameters in one. The baseline state is characterized by the size *S*, the out-connectivity *E* and the betweenness centrality BC of the RA and the seed. The resected network (i.e. the network after the VR of the RA was performed) is characterized by the new out-connectivity *E* and BC of the seed, and by their respective decreases due to the virtual resection, i.e. Δ*E*_seed_ = *E*_seed_(baseline) − *E*_seed_(resection) and similarly ΔBC_seed_ = *E*_seed_(baseline) − BC_seed_(resection). Significant effects are indicated by bold font in the p-value.

| | | Effect | $r^2$ | $p$ |
| --- | --- | --- | --- | --- |
| Model: | $\beta\kappa/\gamma$ | + | 0.09 | **0.01** |
| Baseline: | $S_{\text{RA}}$ | + | 0.33 | **$8 \cdot 10^{-8}$** |
| | $S_{\text{seed}}$ | − | 0.00 | 0.7 |
| | $E_{\text{RA}}$ | + | 0.11 | **0.004** |
| | $E_{\text{seed}}$ | − | 0.02 | 0.2 |
| | $\text{BC}_{\text{RA}}$ | + | 0.08 | **0.0** |
| | $\text{BC}_{\text{seed}}$ | − | 0.05 | 0.06 |
| Resection: | $E_{\text{seed}}$ | − | 0.10 | **0.006** |
| | $\Delta E_{\text{seed}}$ | + | 0.06 | **0.04** |
| | $\text{BC}_{\text{seed}}$ | − | 0.11 | **0.004** |
| | $\Delta \text{BC}_{\text{seed}}$ | + | 0.15 | **$5 \cdot 10^{-4}$** |

The variables considered in the previous analysis are not independent: the different centrality metrics are related, and the properties of the RA also impact the structural effect of the resection, for instance. Therefore, to identify the most relevant model properties determining the effect of the resection, we performed a step-wise linear regression analysis. As dependent variable we considered the normalized effect of the resection, in logarithmic scale, log (*δ*_*IR*_ (*i*, seed_*j*_)), for each patient *i* and seed *j*. The resulting (adjusted) model is shown in figure 8. We found that only three variables survived: the size of the RA, *S*_RA_, the BC of the seed in the resected network, BC_seed,VR_, and the decrease in BC of the seed due to the resection, ΔBC_seed_. The partial effect of all other variables was not significant once these three metrics were included. The adjusted model achieved a goodness-of-fit *r*^2^ = 0.468.

**Figure 8:**
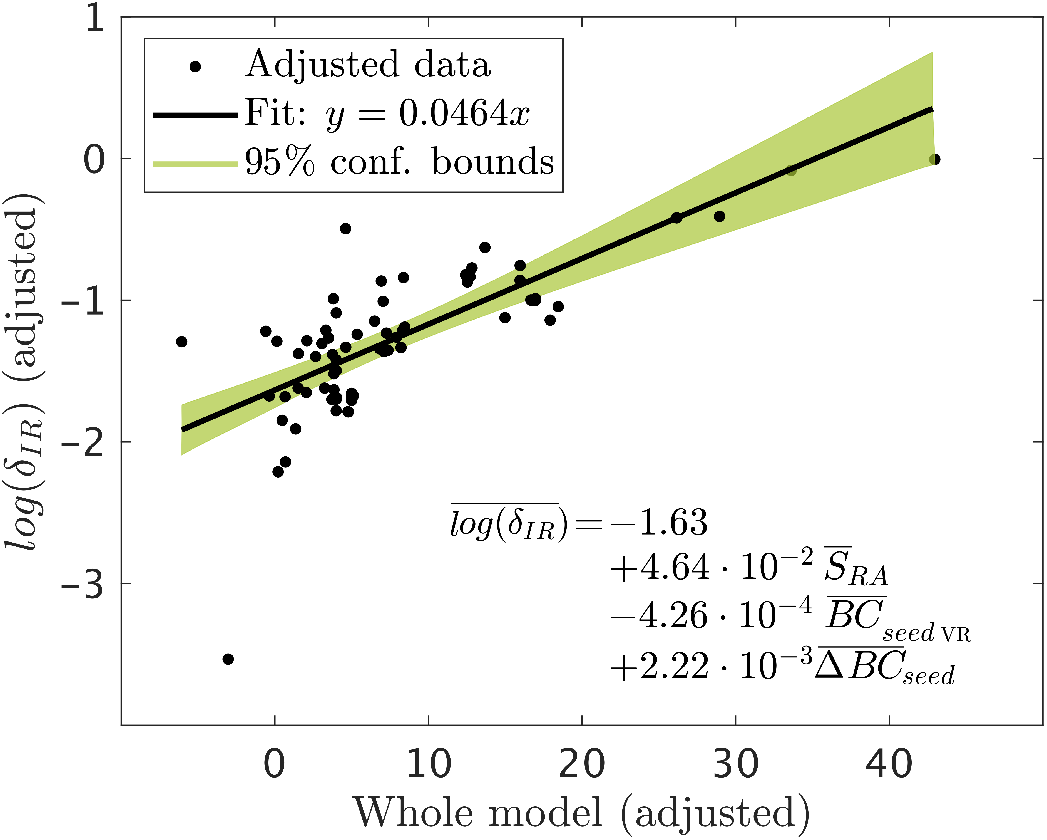
Added variable plot (partial regression leverage plot) of the linear regression model resulted from the step-wise regression analysis. Data-points indicate the adjusted response values against the adjusted predictor variable values, the solid line indicates the adjusted linear fit, and the shaded areas the 95% confidence intervals. The bars over the variable names indicate adjusted variables. The statistical details of the fit are: number of observations = 75, degrees of freedom *df* = 71, root Mean Squared Error *rmse* = 0.397, *r*^2^ = 0.489, Adjusted *r*^2^ = 0.468, F-statistic vs. constant model = 22.7, *p* = 2.1 · 10^10^.

These analyses indicate that the effect of virtual resections in the model is predominantly characterized by the size of the RA, as one might expect, but also by the centrality properties of the seed in relation to the RA. That is, both the hub status of the seed *after* the resection, and the decrease in hub status due to the resection were important for the decrease in spreading, but not the initial hub status *per se*.

## 3 Discussion

We have defined a computational framework to simulate seizure propagation and epilepsy surgery based on epidemic spreading models that integrate patient-specific data. A model was built for each patient based on their individual AEC-MEG brain network to combine structural and functional connectivity, and the propagation of ictal activity over the brain was modelled by means of a simple epidemic spreading model. The model was further individualized for each patient by fitting the main parameters, namely the spreading and recovery rates, and the network density, to the patient-specific seizure propagation pathways, as derived from SEEG data. We found that the model reproduced the main aspects of seizure propagation for all patients, indicating that these simple spreading rules are enough to encode the basic aspects of seizure propagation. Once fitted for each patient, the model can be used to generate alternative hypotheses about the seizure onset zone, or to test the effect of resection strategies, as we have illustrated in this study.

Epidemic spreading models capture the basic mechanisms of processes that propagate on networked systems, and are supported by a well-grounded mathematical and computational framework [51, 56] that we can use to our advantage in the context of epilepsy surgery. For example, the fundamental role of hubs on surgical outcomes is expected from the perspective of epidemic spreading, as the epidemic threshold is known to vanish for networks with a scale-free degree distribution (and therefore high-degree hubs) [51]. On the contrary, a strong community structure can trap the epidemic in one of the communities, preventing large-scale spreading [57, 58], which relates to the clinical observation that seizure propagation can often be restricted to one or a few brain lobes [1], as is the case in focal epilepsy. The fact that epidemic spreading provides a good representation of seizure propagation suggests that other network characteristics that are known to play an important role in epidemic spreading processes, such as temporal changes in connectivity [59–62] due to mal-adaptive plasticity over long time scales (months to years) [63], degree correlations [64] or dimensionality [56, 65, 66], may also affect seizure propagation.

In this study we decided to use MEG networks as the backbone of seizure propagation, in contrast with other studies [30, 33, 37, 44] based on DTI (Diffusion Tensor Imaging) data. In a previous study we showed that the AEC metric, whilst based on functional connectivity, retains information on the structural pathways [34] and can be used as a cost-effective proxy for structural connectivity: DTI is not typically part of the standard pre-surgical evaluation of the patients, has a much higher computational cost than AEC-MEG, and has low sensitivity to long range connections, in particular inter-hemispheric ones [67].

### 3.1 Epidemic spreading predicts surgery outcome

One of the main goals of computational studies of epilepsy surgery is to predict surgery outcome and optimize surgical planning. In our modeling framework, we found that the model, when considering the RA as the epidemic seed, yielded a better fit (as given by the correlation between the modelled and recorded seizures) for SF than for NSF patients, and the difference was significant when considering the population model. Moreover, considering the model fit as a classification parameter led to a good differentiation between the SF and NSF groups, with an AUC of 0.841 for the individual models and 0.886 for the population model, indicating that the goodness-of-fit could be used as a predictor for surgical outcome. This result also suggests an explanation for the different surgical outcome for the SF and NSF groups as, according to the model, the RA was a better approximation to the SOZ for SF patients, and consequently its removal was more likely to lead to seizure freedom, as was indeed the case. Thus, if a better hypothesis on the SOZ could be made for NSF patients using the computational model, then the resection strategy could also be improved, potentially leading to a better outcome.

It is important to notice, however, that other interpretations are possible. The model fit results are dependent on the SEEG sampling, which may have been inadequate for NSF patients, so that relevant aspects of the seizures were missed [44]. In this case, the models would not be able to improve the hypothesis on the SOZ, although a poor fitting result could still be used as an indication that more pre-surgical evaluations are needed, with e.g. alternative spatial sampling. Finally, it is also possible that the worse fit of the model may have been caused by more prevalent non-linear or multi-scale effects for NSF patients, that would make seizure dynamics deviate from a spreading process [68]. In this case the mismatch would not indicate an error in the SEEG sampling or the surgery planning, but point towards an intrinsic difference in seizure dynamics.

In order to shed light on this question, we made use of the seizure model to generate alternative hypotheses on the SOZ by measuring for each individual ROI the likelihood of generating the observed seizures (Fig. 5). At a group level, RA ROIs were significantly more likely to generate the observed seizures than non-RA ROIs, as expected. However, for 8 out of 15 cases RA regions did not show higher seed-likelihood than non-RA regions, and the ROI with the maximum seed-likelihood did not belong to the RA for any case. This suggests that the most likely seeds according to the model were close to the RA, but did not belong to it. This result is in agreement with other modelling studies that found modelled SOZ that did not completely overlap with the resected areas, even for SF patients [28, 31, 37], and is likely associated with the incomplete sampling of the SEEG electrodes. We hypothesize that it may also be related to the finding of pathological hubs whose disconnection from the SOZ can be enough to lead to seizure freedom, even when the SOZ or the pathological hub remain unresected [18, 21, 25, 35].

Remarkably, we also found that the difference in goodness-of-fit between the SF and NSF groups disappeared when considering the optimal single-seed fit. This suggests that one can find alternative hypotheses of the SOZ for NSF patients that lead to a good model fit. Then, a resection targeting these regions might lead to a better outcome, in agreement to our finding that the goodness-of-fit is a good predictor for surgical outcome. However, as this study comprised only 4 NSF patients, more data would be needed to validate this finding.

Finally, we performed a virtual resection analysis to simulate the effect of the resective surgery *in silico* for each patient [31, 33, 34]. We found that virtual resections of the RA led to a significant decrease in seizure propagation. Here we considered the relative decrease in spreading and not seizure extinction to characterize the effect of a resection in the model, as spreading in the model is never null (since the seed is always infected) and the considered VRs seldom disconnected the seed completely. Therefore, the relative decrease is more informative than absolute post-surgery spreading, as it reduces the influence of specific modelling choices. We found that the effect of the resection was predominantly affected by i) the size of the resection, ii) the decrease of betweenness centrality of the seed as a consequence of the resection; and iii) the betweenness centrality of the seed *after* the resection. Remarkably, the centrality of the RA has been associated before with surgery outcomes, with the removal of network hubs being associated with seizure freedom [18, 21, 33]. Here we have found that, more than the centrality of the removed regions, it is the centrality of the SOZ in relation to the RA that will determine the effect of the resection. Given that the gold-standard for the actual SOZ in the clinical setting cannot be known, and this often needs to be approximated by the RA, a direct comparison with clinical results is difficult. Prospective studies that include alternative hypothesis for the SOZ, however, can be used to gain more insight in this regard. Finally, other model properties, such as the spreading-to-recovery ratio, also correlated significantly with the relative decrease in spreading, but the effect did not survive the step-wise linear regression analysis, and was mediated by the size of the resection and the centrality of the seed.

In our study, we found that the relative effect of VRs was larger for the SF +than for the NSF group, and we found AUC values between 0.63 and 0.75 when using the normalized decrease in spreading to classify the patients according to surgical outcome. This again indicates, in agreement with our results based on the goodness-of-fit, that the RA is a better approximation of the SOZ for SF patients, which is information that can be gained with the model prior to the surgery for a given resection strategy. However, the small sample size in this study limits the predictive power of the results, and the difference in effect between the SF and NSF was not significant. Overall, a larger patient group, including more than one SEEG seizure pattern per patient, would help to improve the predictive power of the model.

### 3.2 Modeling considerations and clinical application

Epilepsy surgery models need to be individualized for each patient if they are to be of clinical use, in order to take into account the patient-specific brain network and seizure dynamics. This presents a transversal problem in the modeling of epilepsy surgery, as individualizing the models requires extensive data, which is not always available. The existing data on seizure spreading is typically based on SEEG recordings which, whilst presenting high temporal and spatial resolution, are limited by sparse spatial sampling, which is known to impact the characterization of the seizures and outcome prediction [69], and can lead to bias in the results [46]. Here we have considered the role of the whole brain network in seizure spreading by using a whole brain atlas and MEG data, to reduce the bias due to sparse sampling, but even then only the regions sampled by the SEEG electrodes could be taken into account to fit the model.

In order to simplify the modelling framework, in this study we considered a simple spreading model as the basis for seizure propagation, but there were still specific limitations associated with the modeling scheme. In particular, the propagation of ictal activity captured by the SEEG electrodes is not a binary process, as it was assumed here. On the contrary, ictal activity presents in different qualitative and quantitative forms, and the reduction of the seizure propagation dynamics to a binary activation-inactivation sequence is an oversimplification. Moreover, in order to avoid introducing arbitrary time-scales in the model, the seizure patterns only considered the activation order of the ROIs, and not the activation times, which reduces the resolution of the pattern further (as it cannot distinguish fast from slow spreading).

Despite the low dimensionality of the model, there was still noise in the fitting method. This noise is intrinsic to the limited clinical data and fitting method, and it is not due to the stochastic nature of the SIR dynamics, which was already taken into consideration: the SIR dynamics were run 10^4^ times per iteration, and 10 iterations were performed, and averaged, for each set of parameters. Moreover, the total correlation metric defined by Eq. 2 also takes into account the stochastic nature of the dynamics by weighting every node by its probability of activation in the model. The *parameter noise* can lead to a noisy parameter landscape, with several local maxima. As a consequence, there is the risk of over-fitting the individual models, and only limited information can be extracted from the fitting parameters (i.e. from the values of *κ, β* and *γ* leading to the best individual fits).

In light of these considerations, it is even more important to refine and simplify the modelling frameworks so as to minimize over-fitting problems and improve the generalizability of the models. A deeper understanding of the biophysical mechanisms leading to seizure generation and propagation will help reducing the number of model parameters that need to be fitted numerically. For instance, considering spreading models, one could systematically –over a large enough patient cohort– study whether the spreading dynamics of epilepsy patients is poised in the supercritical regime and it moves to the sub-critical regime after the surgery. This general information could then be integrated into the model to simplify the fitting algorithm.

### 3.3 Population model

The population model was defined here as the model (as defined by the set of control parameters) leading to the best average fit over the patient group. Despite its name, this model is still individualized for each patient: it considers the patient-specific network (including the link weights that define the local spreading probabilities) and seed regions. As it is shown in figure 4, the resulting fit diagram displays the familiar behavior of a phase transition, with an intermediate region of high correlation (good population fit), separating regions of low correlation (poor population fit). Remarkably, most individual fit points were located in this intermediate region, with the exception of three patients who presented very “bulky” activation patterns (by visual inspection, not shown), i.e. in which several ROIs got infected simultaneously. More studies with improved data resolution and larger patients cohorts should be able to establish whether there are actual differences in the dynamical repertoire of these two types of patients.

The average model fit achieved by the population model at its optimal point was much smaller (about 1*/*3 on average) than those obtained with the individual models. However, the difference between the SF and NSF groups not only still held in this model, but it became stronger and significant 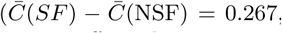, *p* = 0.02 and *AUC* = 0.886). This suggests that the loss of detail in the fitting does not affect the main aspects of seizure propagation, and signals towards the possible over-fitting of the individual models due to parameter noise. In this case, the population model, even though reducing the overall fit, provided a more reliable description of the system.

A reliable population model would drastically increase the clinical applicability of this framework, as its application would not rely on patient-specific SEEG data. SEEG studies are highly invasive, and are avoided in the pre-surgical evaluation whenever possible. Thus, a computational model that could provide relevant information on surgery outcome without the need to be fitted to patient-specific invasive data would provide a valuable tool. Information from other imaging modalities could potentially also be included, such as ictal EEG recordings of epileptiform abnormalities found in MEG or MRI lesions. This information could be incorporated into the model for example as factors that affect the seed-likelihood of the involved ROIs.

### 3.4 Limitations

As we have discussed above, modeling of seizures presents inherent limitations associated with the choices of the dynamical model and fitting procedures [46, 48]. In our case, this translated into difficulties defining alternative seeds and characterizing virtual resections. Identifying seed regions dramatically increases the dimensionality of the fitting problem, even when considering linear approximations as we have done here. Estimating seed-probability maps for seeds of increasing sizes becomes a combinatorial problem that soon loses tractability. The use of optimization algorithms (such as simulated annealing, genetic algorithms or deep learning, for instance [33, 34]) would reduce the computational burden but still be affected by an exponentially larger number of local maxima as the seed size increases, due to the parameter noise. In our analysis we have opted for a linearization approach, characterizing the effect of single seeds and following a recursive method to derive seeds of increasing sizes. Deriving more detailed seeds would require larger amounts data, for instance by considering several seizures per patient.

The use of SEEG data to fit the model posses another limitation for its clinical use, given that SEEG recordings are highly invasive and not always part of the presurgical evaluation. A reliable population model (that can be individualized for each patient by considering their individual brain connectivity) would allow us to also use the model for patients without SEEG recordings, as we have discussed above. Moreover, as we have shown in this study, such a model can also help reducing parameter noise, leading to more robust results than the individual models.

Another main limitation of the study is the small size of the patient group, which complicates the validation of the results. For instance, the VR analysis points towards a larger decrease in seizure propagation after the virtual resections for the SF group, but the difference is not significant. A larger cohort would allow us to improve the classification analysis to clarify this finding. Moreover, increasing the patient group size would also improve the formulation of the population model.

Modeling of virtual resections suffers from some inherent limitations we well. First of all, virtual resections are typically modelled by removing or disconnecting nodes or links from the network [1, 30, 70]. However, this does not account for the generalized effect that a local resection can have on the network [71] nor does it consider plasticity mechanisms [8, 61, 72–74] which are known to occur following brain lesions and resections [75, 76]. An even more fundamental limitation is the difficulty of the validation of the results, as different resection strategies cannot be tested clinically. Validation must always be done indirectly, by comparing the model predictions (regarding for instance the location of the SOZ or surgery outcome) with the clinical results [28, 29, 31, 33, 34]. In this work, we have made use of multi-modal patient-specific data to optimise and validate the model and, as final validation mechanism, we have considered surgery outcome. This can only be the first step, however, as the ultimate goal is to use the computational models to aid epilepsy surgery planning. Prospective or pseudo-prospective studies in which the models are used before or without knowledge of the surgery to predict outcome at an individual level (i.e. not only at a group level) will be necessary in the future to test the applicability of the model on a clinical setting.

## 4 Conclusion and outlook

Epidemic spreading models fitted with patient-specific data reproduce the individual seizure propagation patterns. This simple framework is sufficient to encode the fundamental aspects of seizure propagation on brain networks. Our results highlight that such individualized computational models may aid epilepsy surgery planning by identifying alternative seed regions and/or resection strategies, with the ultimate goal of improving surgery outcome rates.

## 5 Methods

### 5.1 Patient group

We retrospectively analyzed 15 patients (9 females) with refractory epilepsy. All patients had undergone resective surgery for epilepsy at the Amsterdam University Medical Center, location VUmc, between 2016 and 2019. All patients had received a magnetoencephalography (MEG) recording, had undergone an SEEG (stereo-electroencephalography) study, including post-implantation CT-scans, and underwent pre- and post-surgical magnetic resonance imaging (MRI). All patients gave written informed consent and the study was performed in accordance with the Declaration of Helsinki and approved by the VUmc Medical Ethics Committee.

The patient group was heterogeneous with temporal and extratemporal resection locations and different etiology (see Table 2 for details). Surgical outcome was classified according to the Engel classification at least one year after the operation [77]. Patients with Engel class 1A were labelled as seizure free (SF), and patients with any other class were labelled as non seizure free (NSF). 4 patients were deemed NSF.

**Table 2:** Patient data. Ep. = Epilepsy, y = years, *S*_=_ number of resected ROIs, #E = number of intracranial electrodes, #ECP = total number of electrode contact points, *N*_*SR*_ = number of BNA ROIs sampled by the SEEG electrodes. F = female, M = male, R = right, L = left.

| Case | Sex | Resected Area | $S$ | Engel Score | #E | #ECP | $N_{SR}$ |
| --- | --- | --- | --- | --- | --- | --- | --- |
| P1 | F | R Frontal | 4 | 1A | 13 | 128 | 47 |
| P2 | F | R Temporal, Occipital | 13 | 1A | 14 | 142 | 50 |
| P3 | F | L Temporal, Occipital | 5 | 1A | 15 | 144 | 53 |
| P4 | M | R Temporal | 13 | 1A | 13 | 126 | 49 |
| P5 | F | R Temporal | 10 | 1A | 11 | 109 | 42 |
| **P6** | **F** | **R Lat. Temporal** | **5** | **2A** | **9** | **99** | **40** |
| P7 | F | L Temporal | 5 | 1A | 11 | 110 | 44 |
| P8 | F | L Parietal | 4 | 1A | 10 | 104 | 37 |
| P9 | M | R Post. Lat. Temporal,<br>Post. Insula, Post. Parietal | 3 | 1A | 12 | 102 | 38 |
| **P10** | **F** | **R Temporal** | **13** | **2D** | **11** | **114** | **45** |
| **P11** | **F** | **L Frontal** | **4** | **2C** | **13** | **117** | **47** |
| P12 | M | L Frontal | 6 | 1A | 12 | 124 | 40 |
| P13 | M | L Temporal | 5 | 1A | 12 | 106 | 30 |
| **P14** | **F** | **L Temporal** | **6** | **3A** | **15** | **194** | **60** |
| P15 | M | R Temporal | 12 | 1A | 10 | 107 | 32 |

### 5.2 Individualized Brain Networks

The individualized computer model was based on the brain network of each patient, which was reconstructed in the Brainnetome Atlas (BNA) from MEG scans, as follows:

- **Pre-operative MRI scans** were used for co-registration with the MEG data. MRI T1 scans were acquired on a 3T whole-body MR scanner (Discovery MR750, GE Healthcare, Milwaukee, Wisconsin, USA) using an eight-channel phased-array head coil. Anatomical 3D T1-weighted images were obtained with a fast spoiled gradient-recalled echo sequence. During reconstruction, images were interpolated to 1 mm isotropic resolution.
- **MEG recordings** were obtained during routine clinical practice using a whole-head MEG system (Elekta Neuromag Oy, Helsinki, Finalnd) with 306 channels consisting on 102 magnetometers and 204 gradiometers. The patients were in supine position inside a magnetically shielded room (Vacummshmelze GmbH, Hanau, Germany). Typically, three eyes-closed resting-state recordings of 10 to 15 minutes each were acquired and used in the presurgical evaluation for the identification and localization of interictal epileptiform activity. The first of these recordings of sufficient quality was used here to generate the brain network. The data were sampled at 1250 Hz, and filtered with an anti-aliasing filter at 410 Hz and a high-pass filter of 0.1 Hz. The head’s position relative to the MEG sensors was determined using the signals from 4 or 5 head-localization coils that were recorded continuously. The positions of the head-localization coils and the outline of the scalp (roughly 500 points) were measured with a 3D digitizer (Fastrak, Polhemus, Colchester, VT, USA).
- **MEG pre-processing**. The temporal extension of Signal Space Separation (tSSS) [78, 79] was used to remove artifacts using Maxfilter software (Elekta Neuromag, Oy; version 2.1). For a detailed description and parameter settings see Hillebrand et al. [80]. The MEG data were filtered in the broadband (0.5 − 48.0 Hz).
- **MEG & MRI co-registration**. The points on the scalp surface were used for co-registration of the MEG scans with the anatomical MRI of the patient through surface-matching software. A single sphere was fitted to the outline of the scalp and used as a volume conductor model for the beamforming approach.
- **Source reconstruction: beamforming**. Neuronal activity was reconstructed using an atlas-based beamforming approach, modified from Hillebrand et al. [81], to reconstruct the time-series of neuronal activation of the ROI centroids [82]. We considered the 246 ROIs of the BNA atlas [83], whose centroids were inversely transformed to the co-registered MRI of the patient. Then, a scalar beamformer (Elekta Neuromag Oy; beamformer; version 2.2.10) was applied to reconstruct each centroid’s time-series, as detailed elsewhere [82].
- **Processing**. The time-series of each centroid were visually inspected for epileptiform activity and artifacts. On average, 58 ± 11 interictal and artefact-free epochs of 16384 samples were selected for each patient. The epochs were further analyzed in Brainwave (version 0.9.151.5 [84]) and were down-sampled to 312 Hz, and filtered in the broadband (0.5 - 48 Hz).
- **Functional networks** were generated considering each brain region as a node. The elements *w*_*ij*_ of the connectivity matrix, indicating the strength of the connection between ROIs *i* and *j*, were estimated by the AEC (Amplitude Envelop Correlation) [85–88]. The uncorrected AEC (i.e. without correcting for volume conduction) connectivity metric was selected as it maintains information on the structural connectivity pattern, whilst including information on long-range functional connections. AEC values were re-scaled between 0 (perfect anti-correlation) and 1 (perfect correlation), with 0.5 indicating no coupling [89]. Functional networks were thresholded at different levels *θ* indicating the percentage of remaining links in the network, and the resulting average connectivity *κ* of the network, indicating the average number of links that each node had, was determined. Notice that the networks were thresholded but not binearized, so that *w*_*ij*_ takes values between 0 and 1. An exemplary adjacency matrix is shown in figure 1 under “Brain Network”.

### 5.3 Resection Area

The resection area (RA) was determined for each patient from the three-month post-operative MRI. This was coregistered to the pre-operative MRI (used for the MEG co-registration) using FSL FLIRT (version 4.1.6) 12 parameter affine transformation. The resection area was then visually identified and assigned to the corresponding BNA ROIs, namely those for which the centroid had been removed during surgery. An exemplary RA is shown in figure 1 under “RA”.

### 5.4 Individualized Propagation Pattern

All patients underwent stereo-electroencephalography (SEEG) electrode implantation. The number and location of the intracerebral electrodes (Ad-Tech, Medical Instrument Corporation, USA, 10-15 contacts, 1.12 mm electrode diameter, 5 mm intercontact spacing; and DIXIE, 10-19 contacts, 0.8 mm electrode diameter, 2 mm contact length, 1.5 insulator length, 16 − 80.5 insulator spacer length) were planned individually for each patient by the clinical team, based on the location of the hypothesized SOZ and seizure propagation pattern. The number of electrodes per patient (see table 2 for details) varied between 9 and 15 (average = 12.1 ± 1.8) and the total number of contacts between 194 and 99 (average = 121 ± 24).

The locations of the SEEG contact points (CPs) were obtained for each patient from the post-implantation CT scan (containing the SEEG electrodes) that was co-registered to the preoperative MRI scan using FSL FLIRT (version 4.1.6) 12 parameter afine transformation. Each electrode CP was assigned the location of the nearest ROI centroid. Because BNA ROIs are in general larger than the separation between contact points, different CPs can have the same ROI assigned.

The activation time of each sampled ROI was determined according to the SEEG recording as follows. First, the onset time of ictal activity was identified for each SEEG channel by a clinician expert. Then, the CPs were grouped into activation steps and a seizure pattern was built from one typical seizure for each patient. This activation pattern was then translated into the BNA space, so that the each sampled ROI *i* was assigned an activation step. This constituted the *SEEG seizure pattern*.

### 5.5 Seizure Propagation Model

#### 5.5.1 SIR Dynamics

Seizure propagation was modelled using the Susceptible-Infected-Recovered (SIR) model [51]. Simulation of the epidemic spreading process on the network took place as follows. Initially, all nodes were set in the susceptible state *S*, except for a set of *seed* nodes in the infected state *I*. At each subsequent step, each infected node could propagate the infection to any of its neighbours with probability *βw*_*ij*_, where *β* characterizes the global spreading rate and *w*_*ij*_ the link weight as defined above. Each infected node had a probability *γ* of recovering to the *R* state. Depending on the network structure, the epidemics can show different spatio-temporal spreading profiles described by the probability *p*_*i*_(*t*) that each ROI *i* becomes infected at step *t*. SIR dynamics was simulated in custom-made Matlab algorithms using Monte-Carlo methods, with *N*_*R*_ = 10^4^ iterations of the algorithm for each configuration to assure convergence.

#### 5.5.2 Individualized Propagation Model

The seizure propagation model was fitted for each patient by comparing the spatio-temporal propagation pattern in the model to the patient’s clinical seizure pattern (constructed as described above), when the RA was used as the seed for epidemic spreading. The mean activation time for each ROI was calculated as 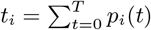, where *T* is the maximum integration time. *t*_*i*_ described the activation sequence of the ROIs during a modelled seizure. By sorting the ROIs according to their mean activation time, we defined the SIR spreading pattern, which indicated the activation order of the involved ROIs. Given that not all BNA ROIs were sampled by the SEEG electrodes, *t*_*i*_ was sub-sampled to the sampled ROIs. This constituted the *SIR seizure pattern*, as shown in figure 1 (under “SIR Seizure Pattern”), which could be compared with the SEEG seizure pattern.

The *goodness of fit* of the model was estimated by taking into account two factors, as illustrated in 2. The first one is the correlation between activation orders of ROIs that became infected both in the SEEG and SIR patterns (see 2A). In order to take into account the noisy nature of the SIR dynamics, such that not the same ROIs get infected in each realization, we considered the weighted Pearson’s correlation coefficient, *C*_*w*_. As correlation weights we used the fraction of realizations that each ROI *i* got infected during a modelled seizure, *P*_*IR*_(*i*). Thus, ROIs that were consistently involved in the spreading weighted more in the correlation than ROIs that were only rarely involved.

The second factor *P*_overlap_ (see 2B) computed the overlap between active and inactive ROIs in the two patterns, to also take into account the actual extension of the seizures, i.e.

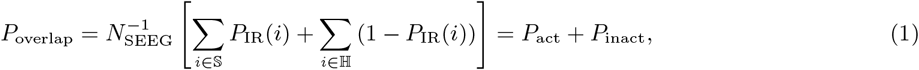

where *N*_SEEG_ is the number of ROIs sampled by the SEEG electrodes (on average = 43.6 ± 7.9), and 𝕊 and ℍ are respectively the sets of active (in the seizure state) and inactive (in the healthy state) ROIs in the SEEG pattern.

Thus, the *total correlation* between the two patterns was defined as

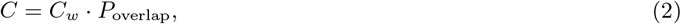

This metric equals 1 in case of exactly equal activation patterns, 0 in the case of null-overlap or correlation, and −1 in the case of complete anti-correlation of activation times (but equal seizure areas). We note however that *C* decays from 1 faster than a simple correlation metric when there are discrepancies between the patterns, since it takes into consideration not only the activation times, but also the activation areas.

In order to fit the model to the SEEG data, the RA was set as the seed of the epidemics, and the model parameters (*κ, β, γ*) were fitted to the data by maximizing *C*, independently for each patient. In order to do so, the SIR dynamics was simulated for a range of values of the free parameters (*β, γ* ∈ {10^−4^, 10^−3^, 10^−2^, 10^−1^}, *κ/N* = {0.025, 0.05, 0.10, 0.20, 0.30}), leading to the 3-dimensional fit-map *C* (*κ, β, γ*). Two exemplary fit-maps are shown in Supp. Fig. 1. In order to minimize noise effects and be able to estimate the error in the measure, *C* values were averaged over 10 iterations of the model (each comprising 10^4^ repetitions for the SIR dynamics), and the error in the measure was defined as the standard deviation across these 10 iterations.

#### 5.5.3 Population model

We defined the population model as the model that provided the best fit overall, by averaging the fit results of all patients. Given that spreading in the model is strongly influenced by the connectivity of the seed, we defined a re-scaled spreading rate, *β*_*RS*_ = *βE*(seed), where *E*(seed) = ∑_*i*∈seed_ ∑_*j*∈nseed_ *w*_*ij*_ is the total link weight from the seed to the rest of the network, and nseed is the set of nodes that do not belong to the seed. For a given *β, E*(seed) characterizes the expected number of infected regions in the first steps of the seizure spreading. Thus, *β*_*RS*_ characterizes the actual spreading probability of the seizure in the model, combining in a single parameter *κ* and *β*, and also taking into account the different seed sizes. Therefore, in order to build the population model, we expressed the individual fit results in terms of *C*(*β*_*RS*_, *γ*), and averaged the individual fit maps to obtain the population fit 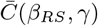.

#### 5.5.4 Alternative seed regions

Alternative seed regions were found in the model by considering each ROI *R* as the single seed of epidemic spreading, once the model was fitted to the patient-specific seizure propagation patterns. Then, the seed-likelihood of the ROI was defined as the total correlation between the SEEG and SIR patterns, *C*_*R*_. Only ROIs leading to spreading were included in the analyses. From this analysis we estimated the best fit given by a single ROI, referred to as *“Best”* and the average value of the fit given by the RA regions, when considered individually as seeds, referred to as ⟨*RA*⟩. Finally, for comparison purposes we also estimated the average model fit given by random seeds (*N* = 20) of the same size of the RA, referred to as *RND*.

### 5.6 Simulation of Resections

We conducted virtual resections (VRs) of the ROIs that were part of the RA to simulate the effect of the surgery in the model. In order to do this, the nodes belonging to the RA were disconnected from the network by setting to 0 all their connections. The effect of each resection was characterized by the normalized decrease in spreading in the resected network (VR) with respect to the original (or baseline, BS) spreading: *δ*_VR_ = (IR_BS_ − IR_VR_)*/*IR_BS_, where *IR* is the fraction of nodes that became infected at any point during the modelled seizure, that is, *IR* = *I*(*t* → ∞) + *R*(*t* → ∞).

The model parameters were chosen as the optimal fitting parameters for each patient, whereas the seed regions were defined following a recursive optimization method. Starting from the best single seed, all possible combinations of this node with the remaining 245 nodes were tested as spreading seeds, and the one leading to the best model fit (i.e. maximum total correlation) was chosen. This process was subsequently iterated until seeds of size 5 were obtained. To account for the large differences in seed sizes and connectivity, we re-scaled the spreading rate by the fraction between the out-connectivity (number of links to the rest of the network) of the RA and the considered seed.

To understand what network and model characteristics relate to the effect of the resection on spreading we estimated the Pearson correlation coefficient between the normalized decrease in spreading due to the resection, *δ*_VR_, and different network and model metrics. In particular, as model metric we considered the spreading-to-recovery ratio, *βκ/γ*, which takes into account that spreading is enhanced by *β* and *κ* and slowed by *γ*. As network metrics we considered the size *S* (number of nodes), out-connectivity *E* (number of links to the rest of the network) and average betweennees centrality BC of the RA and the seed. For the seed we considered three scenarios: the baseline level (BS, prior to the resection), the post-resection level (VR) and the decrease due to the resection, Δ.

In order to identify the most relevant model variables to predict the effect of a virtual resection in the model, we performed a step-wise linear regression model analysis. We used the *Statistical and Machine Learning Toolbox* with standard settings (*stepwiselm* function with default settings). Only linear effects, and no interaction effects, were allowed in the linear model. As dependent variable we considered the normalized effect of the resection in logarithmic scale, log (*δ*_*IR*_). As independent variables we considered the same network and model metrics as in the pairwise correlation analyses.

### 5.7 Statistics

The weighted correlation coefficient was used to determine the correlation between the SEEG and SIR seizure propagation patterns. For comparisons between resected and non-resected areas, and between different seed definitions, we used paired Student’s t-tests, whereas for comparisons between SF and NSF patients, we used unpaired Student’s t-tests. Significance thresholds for statistical comparisons were set at *p <* 0.05.

We performed a receiver-operating characteristic (ROC) curve analysis to study the patient classification based on the goodness of fit of the models and the normalized effect of virtual resections. A positive result was defined as good (SF) outcome.

In order to account for the noise in the SIR model, the dynamics were run 10^4^ to attain each SIR seizure pattern, and this was repeated 10 times to obtain averaged values. The errors were defined as the standard deviation between the 10 realizations of the model. The same procedure was used in the virtual resection analysis.

### 5.8 Data availability

The data used for this manuscript are not publicly available because the patients did not consent for the sharing of their clinically obtained data. Requests to access to the data-sets should be directed to the corresponding author. All user-developed codes are publicly available in GitHub.

## Data Availability

The data used for this manuscript are not publicly available because the patients did not consent for the sharing of their clinically obtained data. Requests to access to the datasets should be directed to the corresponding author. All user-developed codes are available from the corresponding author upon reasonable request.

## 6 Acknowledgements

Ana P. Millán and Ida A. Nissen were supported by ZonMw and the Dutch Epilepsy Foundation, project number 95105006. The funding sources had no role in study design, data collection and analysis, interpretation of results, decision to publish, or preparation of the manuscript.

## 8 Author Contributions

A.P.M., E.C.W.S., C.J.S., I.A.N, A.H. conceptualized the study, E.C.W.S., C.J.S., I.A.N, S.I., J.C.B., P.V.M., A.H. participated in the funding acquisition, A.P.M, E.C.W.S., C.J.S, A.H. devised the Methodology, A.P.M. performed the formal analysis, A.P.M, I.A.N, A.H. devised the software and visualization E.C.W.S., C.J.S., P.V.M., A.H. participated in the supervision, E.C.W.S., S.I., J.C.B. provided resources, A.P.M. wrote the original draft and all authors participated in writing, review and editing.

