## Supplementary Information for "The role of epidemic spreading in seizure dynamics and epilepsy surgery"

[1]Amsterdam UMC location Vrije Universiteit Amsterdam, Department of Clinical Neurophysiology and MEG Center, De Boelelaan 1117, Amsterdam, The Netherlands
[2]Amsterdam UMC, Vrije Universiteit Amsterdam, Department of Neurosurgery, De Boelelaan 1117, Amsterdam, The Netherlands
[3]Amsterdam Neuroscience, Brain Imaging, Amsterdam, The Netherlands
[4]Amsterdam Neuroscience, Systems & Network Neurosciences, Amsterdam, The Netherlands
[5]Amsterdam Neuroscience, Neurodegeneration, Amsterdam, The Netherlands
[6]Amsterdam Neuroscience, Cancer Biology and Immonology, Amsterdam, The Netherlands
[7]Amsterdam Neuroscience, Imaging and Biomarkers, Amsterdam, The Netherlands
[8]Faculty of Electrical Engineering, Mathematics and Computer Science, Delft University of Technology, Delft, The Netherlands

#### 1 Model fitting

Figure $S1$ shows the fit results for two exemplary cases, as indicated in the caption. Best fit parameters (used during the analysis) are indicated by full circles.

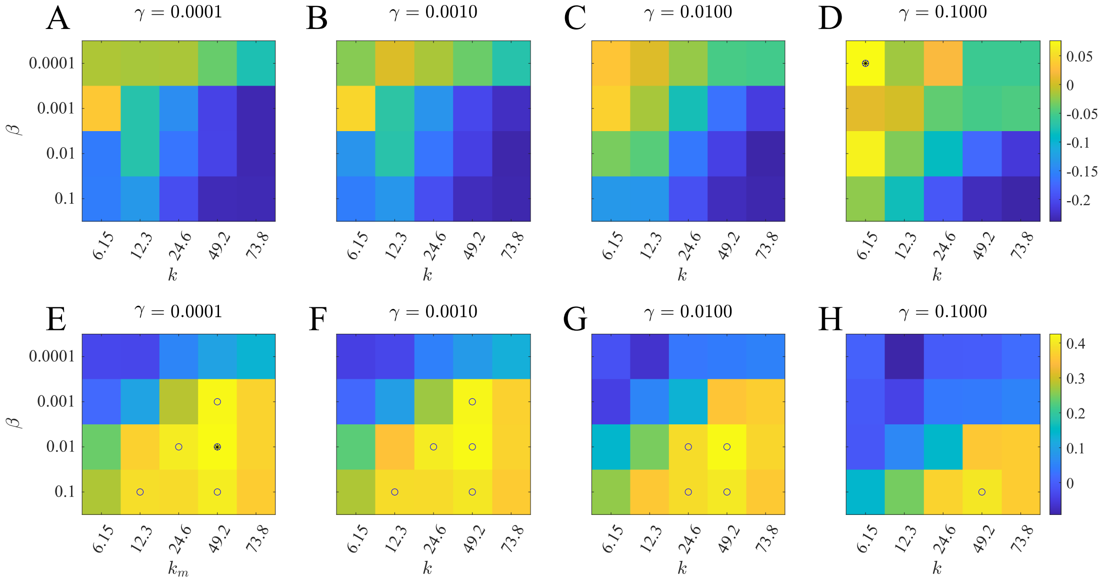


Figure S1: Model fit. Fit results for two exemplary patients (cases 10 and 15, respectively on the top and bottom rows) as given by $C(\beta, \kappa)$ for different values of $\gamma$. The full circle marker indicates the best fit, and empty markers indicate fit points with at least 90% of the maximum value.

### 2 Population model

For the definition of the population model, the individual fit maps are expressed according to the re-scaled (RS) spreading rate $\beta_{RS} = \beta E(\text{seed})$, where $E(\text{seed}) = \sum_{i \in \text{seed}} \sum_{j \in \text{nseed}} w_{ij}$ is the total link weight from the seed to the rest of the network, and nseed is the set of nodes that do not belong to the seed. This integrates in one parameter the effect the global spreading rate, the density of connections, and the size of the seed. Then, the maps for all patients are averaged to create the average population map, as shown in figure $S2$A. The variation across the population is measured via the standard deviation among the individual maps ($S2$B), and the signal to noise ratio is defined as the ratio of the average fit values over the standard deviation, for each set of parameters ($S2$C).

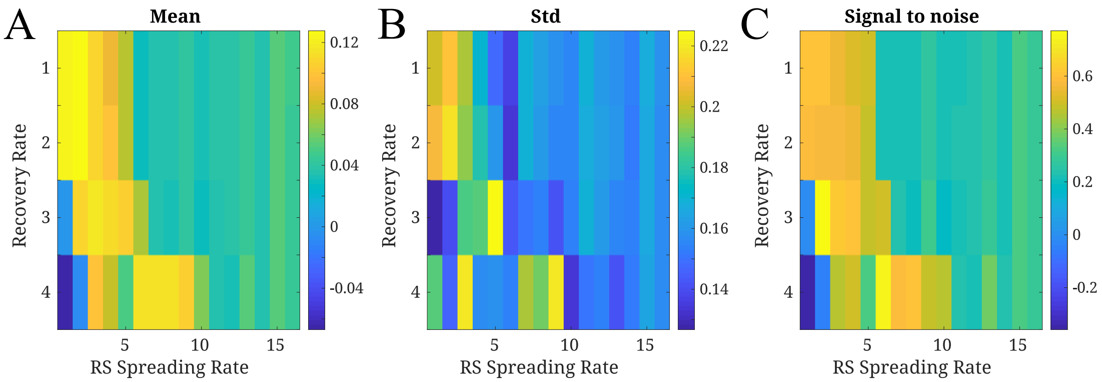


Figure S2: Population model. **A** Average fit map in terms of the recovery rate $\gamma$ and the re-scaled (RS) spreading rate, $\beta_{RS}$. Yellow colors mark high average correlation values. **B** Standard deviation accross the population. **C** Signal to noise ratio given by the ratio between the average values and the standard deviation.

### 3 Alternative seizure onset zones

In this section we report on the details of the statistical analyses in section 2.4: "Alternative Seizure Onset Zones" of the main text. Table $S1$ corresponds to the comparison between the seed-likelihood of resected (RA) and non-resected (NRA) areas. All results correspond to un-paired t-tests. In table $S2$ we report the results from the statistical comparisons between different seed definitions.

| Case | $C_{\text{RA}}$ | $C_{\text{NRA}}$ | Diff. | $t$ | $p$ | $df$ | Sig. |
|---|---|---|---|---|---|---|---|
| 1 | 0.300 | 0.036 | 0.261 | 1.744 | 0.08 | 89 | * |
| 2 | 0.173 | −0.018 | 0.191 | 2.73 | 0.07 | 186 | * |
| 3 | −0.205 | 0.0186 | −0.223 | −1.10 | 0.22 | 27 | |
| 4 | −0.005 | −0.001 | −0.003 | −0.32 | 0.75 | 193 | |
| 5 | −0.005 | 0.039 | −0.044 | −0.81 | 0.42 | 162 | |
| **6** | 0.067 | 0.029 | 0.038 | 1.48 | 0.14 | 239 | |
| 7 | 0.077 | −0.008 | 0.085 | 1.76 | 0.08 | 242 | * |
| 8 | 0.0233 | −0.007 | 0.030 | 0.59 | 0.56 | 161 | |
| 9 | 0.010 | −0.198 | 0.208 | 5.56 | $< 10^{-4}$ | 244 | ** |
| **10** | −0.016 | −0.003 | −0.013 | −0.41 | 0.69 | 58 | |
| **11** | 0.173 | −0.038 | 0.211 | 4.01 | 0.001 | 244 | ** |
| 12 | 0.340 | 0.100 | 0.240 | 0.63 | 0.54 | 10 | |
| 13 | 0.050 | −0.111 | 0.161 | 8.92 | $< 10^{-4}$ | 244 | ** |
| **14** | 0.062 | 0.051 | 0.011 | 0.06 | 0.96 | 123 | |
| 15 | 0.223 | 0.133 | 0.090 | 2.47 | 0.01 | 244 | ** |

Table S1: Seed likelihood of resected and non-resected areas. Only ROIs leading to non-null spreading are included in the analysis.

| | Diff. | $t$ | $p$ | Sig. |
|---|---|---|---|---|
| Best vs RA | 0.061 | 1.45 | 0.17 | |
| Best vs $\langle$RA$\rangle$ | 0.281 | 6.70 | $< 10^{-4}$ | * |
| Best vs RND | 0.341 | 7.44 | $< 10^{-5}$ | * |
| RA vs $\langle$RA$\rangle$ | 0.220 | 5.47 | $< 10^{-4}$ | * |
| RA vs RND | 0.280 | 4.62 | $3 \cdot 10^{-4}$ | * |
| $\langle$RA$\rangle$ vs RND | 0.060 | 1.70 | 0.11 | |

Table S2: Comparison of the goodness of fit with different seed definitions. Best stands for the best individual seed, RA for the resected area, $\langle$RA$\rangle$ for the average fit of the RA ROIs considered as individual seeds, and RND for random resections of the same size as the RA. To perform the comparison we used a paired t-test between the individual patient fits. $df = 14$ in all cases.

### 4 Virtual resection analysis

In this section we report on the details of the statistical analyses in section 2.5: "Virtual resection analysis" of the main text. Table $S3$ corresponds to the comparison of the effect of VRs of increasing size between the SF and NSF groups (corresponding to the analysis shown in figure 7 of the text). The data corresponding to this analyses are shown in figure $S3$, where each panel corresponds to a different seed size. Finally, table $S4$ indicates the statistical details of the step-wise linear regression analysis (table 1 and figure 8 of the main text).

| $S$ | SF | NSF | diff | $t$ | $p$ |
|---|---|---|---|---|---|
| 1 | −1.13 | −1.87 | 0.740 | 1.61 | 0.13 |
| 2 | −1.02 | −1.44 | 0.414 | 1.48 | 0.16 |
| 3 | −1.12 | −1.57 | 0.448 | 1.62 | 0.12 |
| 4 | −1.24 | −1.55 | 0.314 | 1.34 | 0.2 |
| 5 | −1.25 | −1.45 | 0.200 | 0.81 | 0.4 |

Table S3: Comparison of the effect of virtual resections, as given by the normalized decrease in total spreading, $\delta_{\mathrm{VR}} = (IR_{\mathrm{BS}} - IR_{\mathrm{VR}})/\mathrm{IR}_{\mathrm{BS}}$, between the SF and NSF groups, for different seed sizes $S$. We report here (and in figure $S3$) on $\log(\delta_{\mathrm{VR}})$, since it displays better the different amount of spreading for different patients. $df = 13$ in all cases. diff indicates the difference between the groups, $t$ the $t$-statistic and $p$ the corresponding $p$-value. These results correspond to the analysis shown in figure 7 in the main text and in figure $S3$.

| Adj. Variables | Estimate | SE | $t$ | $p$ |
|---|---|---|---|---|
| Intercept | −1.63 | 0.11 | −15.4 | $3 \cdot 10^{-24}$ |
| $S_{\mathrm{RA}}$ | $4.64 \cdot 10^{-2}$ | $1.1 \cdot 10^{-2}$ | 4.26 | $6 \cdot 10^{-5}$ |
| $\mathrm{BC}_{\mathrm{seed,VR}}$ | $-4.26 \cdot 10^{-4}$ | $1.3 \cdot 10^{-4}$ | −3.33 | $1 \cdot 10^{-3}$ |
| $\Delta\mathrm{BC}_{\mathrm{seed}}$ | $2.22 \cdot 10^{-3}$ | $5.1 \cdot 10^{-4}$ | 4.33 | $5 \cdot 10^{-5}$ |

Table S4: Results from the step-wise linear regression analysis. As independent variables we considered the 11 model and network parameters specific in Table 1 of the main text (spreading ratio, centrality metrics of the RA and seed). Only three variables survived: the size of the RA $S_{\mathrm{RA}}$, the BC of the seed in the resected network, $\mathrm{BC}_{\mathrm{seed,VR}}$, and the change in BC of the seed due to the resection, $\Delta\mathrm{BC}_{\mathrm{seed}}$. The coefficients of the fit are indicated in this table, where SE stands for the standard error of the estimate, $t$ is the t-statistic for a test that the coefficient is zero, and $p$ is the corresponding p-value of the t-test. The main statistics of the fit are as reported in the main text (Table 1).

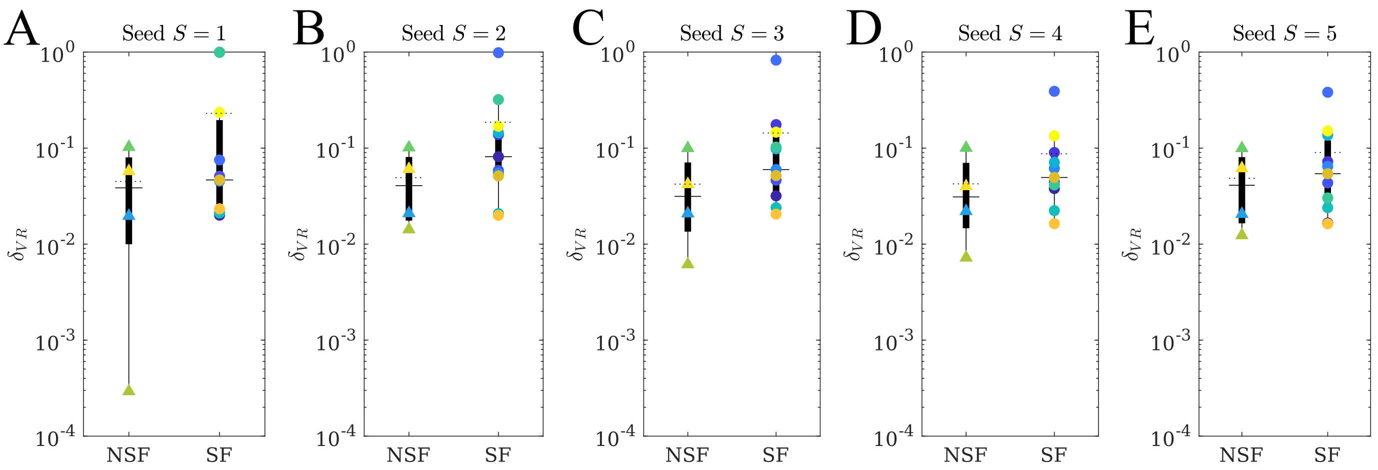


Figure S3: Effect of virtual resections: group effect. Comparison between the normalized decrease in spreading due to the VR of the RA, $\delta_{\mathrm{VR}}$, between the SF and NSF groups. Each panel corresponds to a different seed size as indicated in the header. Mean (median) values are given by dotted (solid) lines. Each data point corresponds to a different patient. The statistical details of the analyses are indicated in table $S3$.
